# Prevalence of metabolic dysfunction-associated fatty liver disease and its association with glycemic control in persons with type 2 diabetes in Africa: a systematic review and meta-analysis

**DOI:** 10.1101/2024.01.02.24300699

**Authors:** Emmanuel Ekpor, Samuel Akyirem, Precious Adade Duodu

## Abstract

Metabolic dysfunction-associated fatty liver (MAFLD) and type 2 diabetes (T2D) are interconnected metabolic disorders that pose serious repercussions on health, yet a comprehensive understanding of the extent of their co-occurrence in Africa is lacking. This study aimed to determine the prevalence of MAFLD and its association with glycemic control (HbA1c) in persons with T2D in Africa. A systematic search was conducted on PubMed, Medline, Embase, Scopus, Global Health, and Web of Science from their inception to December 6, 2023. Data on MAFLD prevalence and correlation coefficients for the association with glycemic control were pooled in random effect meta-analyses. Potential sources of heterogeneity were investigated using subgroup analysis and meta-regression. A total of 10 studies were included in the meta-analysis of MAFLD prevalence, while 2 incorporated in the analysis of the association between MAFLD and glycemic control. The pooled prevalence of MAFLD in persons with T2D was 48.1% (95% CI: 36.1–60.3). By region, the prevalence recorded were 44.7% (95% CI: 28.7–62.0) in sub-Saharan Africa and 55.3% (95% CI: 36.2–73.0) in Northern Africa. We observe an increasing trend in MAFLD prevalence, recording 55.1% (95% CI: 43.6– 66.1) in the recent five years. There was a weak positive correlation between MAFLD and HbA1c (r = 0.33, 95% CI: 0.18 – 0.47). There is a high prevalence of MAFLD in persons with T2D in Africa, with a suggested link between MAFLD and suboptimal glycemic control.

## Introduction

Metabolic Dysfunction-Associated Fatty Liver Disease (MAFLD), formerly referred to as non-alcoholic fatty liver disease (NAFLD), has emerged as a significant public health concern, with a global prevalence of 32·4% [1]. MAFLD is the commonest form of liver disease and it constitutes the majority of liver-related morbidity and mortality cases [2, 3]. The defining features of MAFLD include hepatic steatosis, coupled with the presence of either T2DM, overweight/obesity, or evidence of metabolic dysregulation [4, 5].

T2D is a major risk factor for MAFLD and a predictor of poor liver-related health outcomes among people living with MAFLD [6, 7]. In a previous meta-analysis, researchers found a 55.5% prevalence of MAFLD among individuals with T2D worldwide (compared with the 32.4% in the general population) [7]. Notably, studies indicate that while most individuals with MAFLD do not experience progressive disease, those with concurrent T2D face a two-fold risk of developing advanced liver disease including cirrhosis and advanced fibrosis [8]. Moreover, MAFLD has been linked to an elevated risk of cardiovascular disease, compounding the existing burden in individuals with diabetes [9]. Recent study has reported that among persons at risk for MAFLD, persons with T2D pose nearly a three-fold risk of chronic kidney disease [6]. Even more disconcerting, studies have demonstrated that among individuals with MAFLD, those with T2D are at a higher risk of liver-related mortality compared with those without T2D [10–12].

Given the intricate relationship between T2D and MAFLD, along with its implications for health, clinical practice guidelines recommend screening individuals with diabetes for MAFLD and advanced liver fibrosis [13, 14]. However, evidence supporting this recommendation in Africa is currently limited. The estimated prevalence of MAFLD among individuals with T2D in Africa stands at 30.4%, a figure derived from a synthesis of only four studies [7]. This represents a modest fraction of the total 88 studies considered globally as of September 2018, highlighting substantial gap in our understanding of this crucial health connection in the African context [7]. Africa is experiencing a rapid epidemiological transition, marked by changes in lifestyle, diet, and an increase in the prevalence of metabolic diseases [15]. In Africa, the number of adults living with diabetes was 24 million in 2021 and this figure is projected to experience a substantial increase of 134% by 2024 [16]. Concerningly, the burden of MAFLD is expected to rise in tandem with these changes. This underscores the urgent need for comprehensive research initiatives in the African context to address this health challenge and inform tailored strategies for prevention and management.

The scarcity of data on the epidemiology of MAFLD and T2D in Africa may be a critical limitation in devising region-specific preventive and management strategies. Understanding the prevalence of MAFLD in this demography is crucial, as it can influence healthcare policies, resource allocation, and early intervention efforts. Therefore, this study aimed to determine the prevalence of MAFLD (primary outcome) and its association with glycemic control (HbA1c) in persons with T2D in Africa (secondary outcome).

## Methods

Prior to this study, an extensive search of electronic medical databases was done to ensure no pre-existing systematic review on this study had been conducted. Following this, a protocol for the review was developed and duly registered on PROSPERO (CRD42023491271). The review process adhered to the Preferred Reporting Items for Systematic Reviews and Meta-Analyses (PRISMA) guidelines (**S1 Checklist**) [17].

### Search strategy

A three-step approach was used to identify all relevant studies for this review. Initial search was conducted on PubMed, Medline, Embase, Scopus, Global Health, and Web of Science from their inception to December 6, 2023. These were then supplemented with additional search on Africa-specific databases including African Index Medicus (AIM), and African Journals Online (AJOL). Furthermore, we meticulously examined the reference lists of pertinent studies to ensure a thorough exploration of the available literature. The search strategy for this review was built on the terms “non-alcoholic fatty liver” or “metabolic dysfunction-associated fatty liver”, “type 2 diabetes”, and “Africa”. Controlled vocabulary and keywords to the search terms were used, with the Boolean operators ‘OR’ and ‘AND’ applied appropriately. No limits were applied to the search. The full details of the search strategy are provided in (**S1 Text**).

### Inclusion and exclusion criteria

The inclusion criteria for this review were: 1) observational study design; 2) persons with T2D; 3) studies conducted in an African country; 4) MAFLD prevalence was provided or calculable with enough data provided; 5) involved adult participants aged 18 years or older; and 6) studies published in English language.

Studies were excluded if we were unable to ascertain how MAFLD was diagnosed (e.g., imaging modalities, hepatic steatosis index, liver-spleen attenuation index, fatty liver index, or liver biopsy). However, studies were included irrespective of the absence or presence of secondary liver disease etiologies (such as excessive alcohol consumption) in the T2D participants. This was to enable us to capture studies conducted both before and after the shift from the term NAFLD to MAFLD.

### Study selection and quality assessment

The articles retrieved from our search were imported into Endnote 20 to remove duplicate records. The remaining articles were then uploaded to Rayyan (https://www.rayyan.ai/) for title, abstract, and full-text screening. Relevant information from each study were extracted using a preconceived and standardized data extraction matrix in Microsoft excel. The information retrieved included first author’s name, publication year, country, study design, sample characteristics (sample size, gender distribution, average age of participants, years lived with T2D, average HbA1c, body mass index), and data on MAFLD including the number of T2D participants with the disease and diagnostic tool used. The study selection and extraction of data were executed by two independent reviewers (EE and SA), with any disagreements resolved through discussion with a third reviewer (PAD).

A quality-weighing approach was used to assess the methodological limitations of the included studies. We used the Joanna Briggs Institute (JBI) checklist for prevalence studies [18]. This tool addresses 9 critical questions with response options, “Yes”, “No”, “Unclear” and “Not applicable”. We determined that studies that had 7–9, 5–6, and less than 5 “Yes” were having low, moderate, and high risk of bias respectively.

### Data analysis

A logit transformation of the prevalence data on MAFLD was performed as recommended by Warton and Hui [19], with the 95% confidence interval (CI) calculated using the Clopper-Pearson interval. A DerSimonian-Laird’s random-effect model meta-analysis was used to pool the prevalence of MAFLD. As a secondary outcome, we pooled the correlation coefficients (*r)* across studies to estimate the association between MAFLD and HbA1c. Pooled correlation coefficients were interpreted as weak, moderate, and strong associations for r < 0.4 (or > −0.4), between 0.4 and 0.7 (or −0.4 to −0.7) and > 0.7 (or < −0.7) respectively [20, 21]. We assessed heterogeneity between studies using the I² statistic, with values of 25%, 50%, and 75% indicating low, moderate, and high heterogeneity, respectively [22]. Subgroup analysis and meta-regression were conducted to identify potential sources of heterogeneity, considering variables such as African regions, year of publication, and diagnostic modalities for MAFLD. The presence of publication bias was evaluated using the Egger’s test, with p<0·05 indicating significant publication bias [23].

## Results

A total of 659 references were identified from our search, consisting of 652 records from major databases (PubMed, Medline, Embase, Scopus, Global Health, and Web of Science) and seven from other supplemental searches on AJOL, AIM, and reference lists of relevant articles. Having removed duplicate records, we screened 232 articles based on their titles and abstracts, yielding 29 full-text articles. Ultimately, 10 articles were included in this review (**Figure 1**).

**Figure 1.**
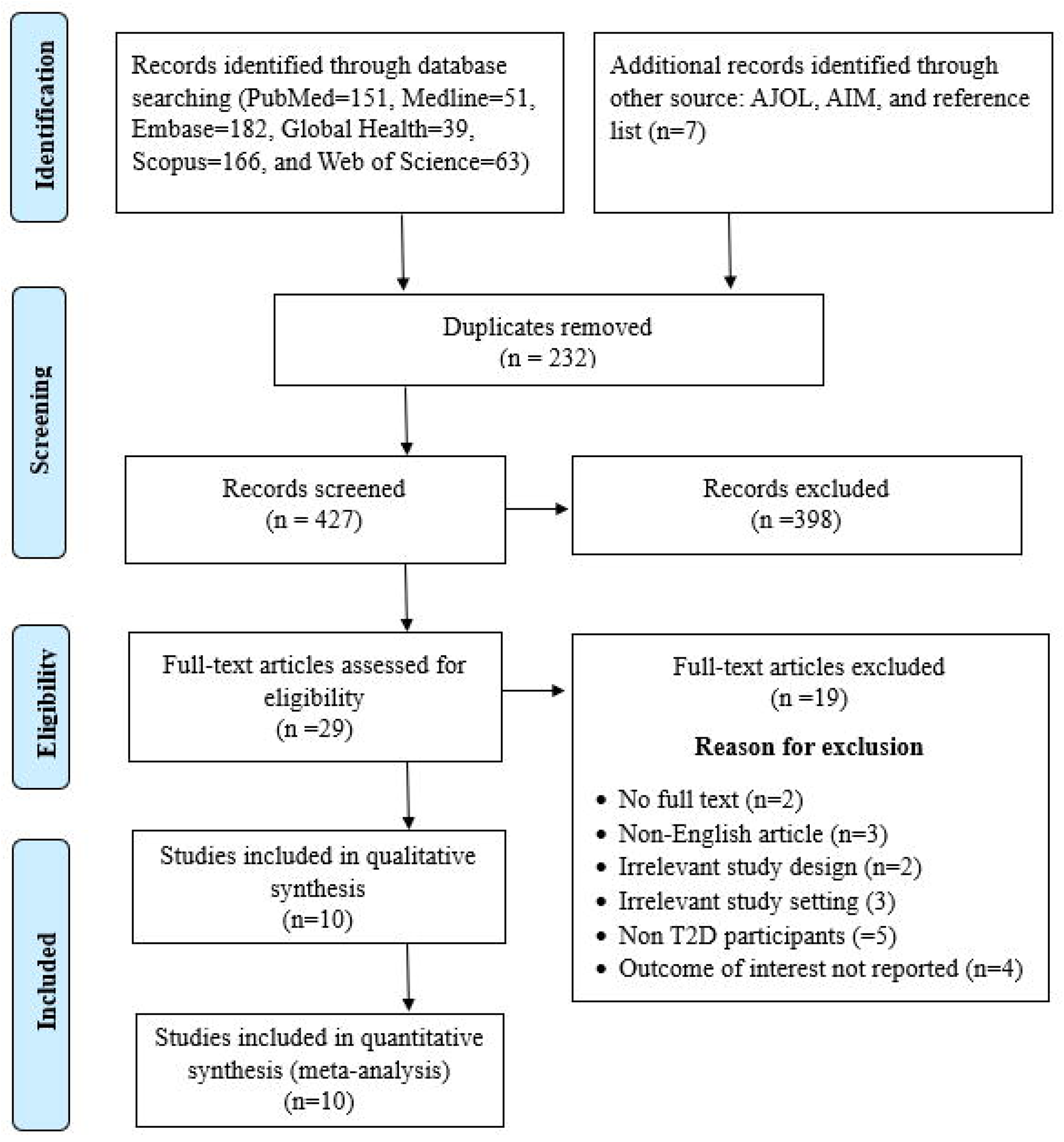
PRISMA flow chart summarizing the article selection process.

### Characteristics of included studies

Full details of the characteristics of the included studies are presented in **Table 1**. This review consisted of nine cross-sectional and one case control studies published between 2011 and 2023. The studies were conducted in 6 different Africa countries, with majority coming from Nigeria (n=3) [24–26], followed by Ethiopia (n=2) [27, 28], Morocco (n=2) [29, 30], and one each from Egypt [31], Ghana [32], and Sudan [33]. A total of 1667 T2D participants were involved, with the sample size ranging from 80 to 281 across studies. Nine studies provided the gender proportion of the participants, with females constituting 61.1%. The participants had a mean age above 50 years and have lived with diabetes for periods ranging from 4 to 10 years. Majority of them were overweight and obese (BMI > 25 kg/m^2^) and had poor glycemic control (HbA1c > 7%). Diagnosis of MAFLD was based mostly on ultrasonography, with just a few using Fatty Liver Index and Hepatic Steatosis Index.

**Table 1.** Characteristics of included studies.

| First author<br>(Year) | Country | Design | Sample<br>size | Female<br>% | Mean<br>age | Mean<br>diabetes<br>duration | Mean<br>HbA1c | Mean<br>BMI | Diagnostic tool<br>of MAFLD | MAFLD<br>prevalence | HbA1c<br>correlation |
| --- | --- | --- | --- | --- | --- | --- | --- | --- | --- | --- | --- |
| Zawdie (2018) | Ethiopia | Cross-sectional | 96 | 53.1 | NR | NR | NR | 23 | Ultrasound | 73 | NR |
| Olusanya (2016) | Nigeria | Case control | 168 | 64.9 | 53.2 | NR | NR | 28.47 | Ultrasound | 16.7 | NR |
| Afolabi (2018) | Nigeria | Cross-sectional | 80 | 62.5 | 60.9 | NR | 8.5 | 25.9 | Ultrasound | 68.8 | $r = 0.270$<br>$p = 0.015$ |
| Abebe (2022) | Ethiopia | Cross-sectional | 101 | 39.6 | NR | 4 | 8 | 25.82 | Fatty Liver Index | 58.4 | $r = 0.35$<br>$p = \square 0.008$ |
| Wiafe (2023) | Ghana | Cross-sectional | 218 | 78.0 | NR | NR | NR | NR | Ultrasound | 51.4 | NR |
| Assarra (2022) | Morocco | Cross-sectional | 180 | NR | 60 | 9.2 | 10.4 | NR | Ultrasound | 45.6 | NR |
| El-Ashmawy (2019) | Egypt | Cross-sectional | 270 | 46.7 | 52.6 | 7.4 | NR | NR | Ultrasound | 73.3 | NR |
| Almobarak (2015) | Sudan | Cross-sectional | 167 | 53.3 | NR | NR | NR | NR | Ultrasound | 50.3 | NR |
| Fennoun (2020) | Morocco | Cross-sectional | 281 | 76.9 | 54.15 | 10.5 | 10.23 | 29.53 | Hepatic Steatosis Index | 45.2 | NR |
| Onyekwere (2011) | Nigeria | Cross-sectional | 106 | 55 | 57.2 | 7.4 | NR | 31 | Ultrasound | 9.5 | NR |

Abbreviations: BMI – body mass index, MAFLD – non-alcoholic fatty liver disease, NR – not reported

### Quality assessment of included studies

Full detail of the quality assessment is shown in (**S1 Table**). The quality score across studies ranged from 8 to 3, with a mean score of 6.1. Majority (n=6) of the studies were of low risk of bias, having a score above 7 [25–28, 30, 32]. In contrast, four studies fell below a cut-off score of 5, indicating a higher risk of bias [24, 29, 31, 33]. Methodological flaws were identified in various areas, including a lack of clarity regarding participant response rates, appropriateness of the sample frame, sampling methodology, adequacy of sample size, and the statistical analysis employed. While the included studies presented prevalence rates and clearly specified the number (n) of T2D patients with MAFLD, none of them provided the confidence interval for the estimates.

### Prevalence of MAFLD

The prevalence of MAFLD ranged from 10.4% (95% CI: 36.1–60.3) in Nigeria [24] to 73.3% (95% CI: 67.6–78.5) in Egypt [31]. The pooled prevalence of MAFLD was 48.1% (95% CI: 36.1–60.3). There was a significantly high heterogeneity among the studies (*I*^2^ = 95%, *p*LJ<LJ0.01) as shown in **Figure 2**.

**Figure 2.**
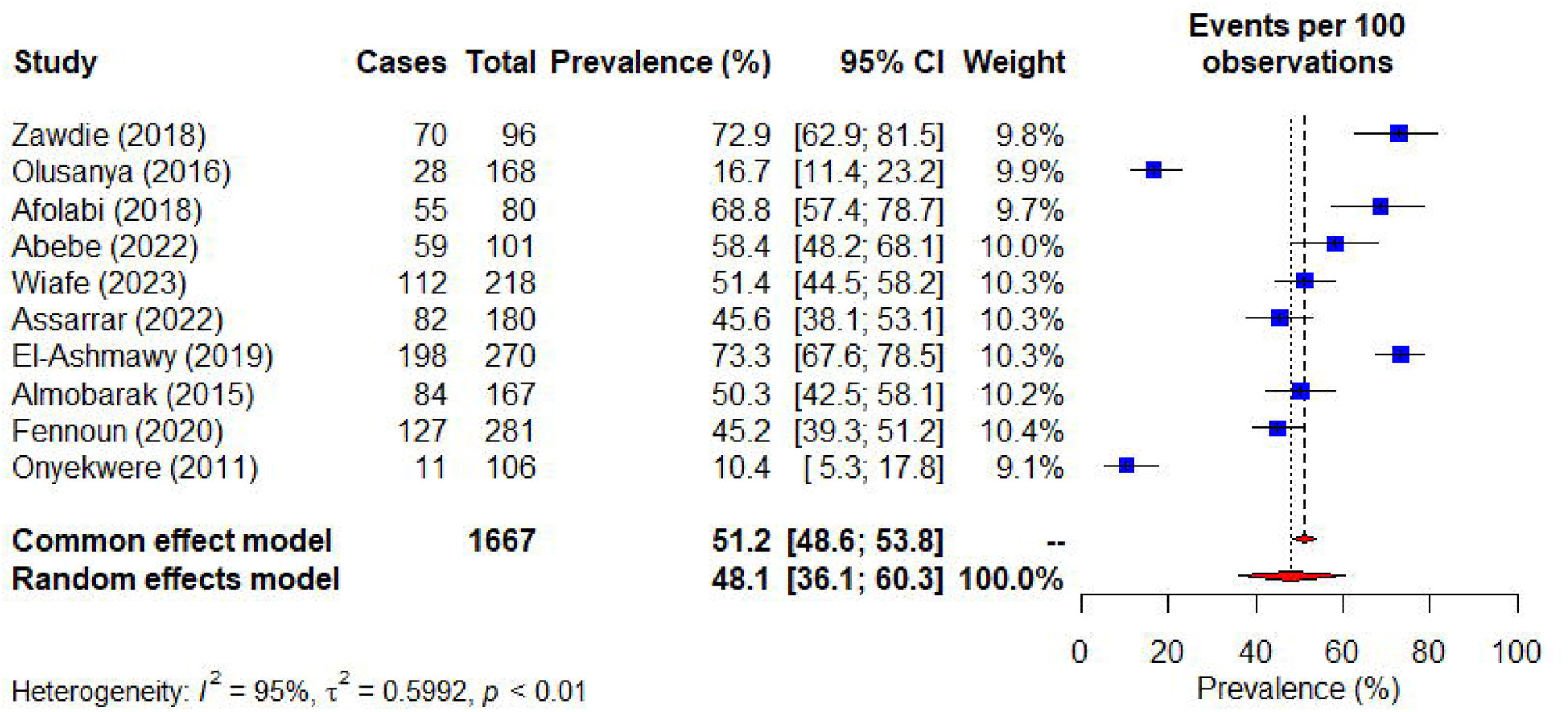
Forest plot for MAFLD prevalence in persons with T2D.

### Subgroup and meta-regression analysis

Subgroup analysis by region revealed a 44.7% (95% CI: 28.7–62.0) prevalence in sub-Saharan Africa and a 55.3% (95% CI: 36.2–73.0) prevalence in Northern Africa. The prevalence of MAFLD according to year of study publication recorded 40.5% (95% CI: 18.6–67.0) in articles published before 2019 and 55.1% (95% CI: 43.6–66.1) for those published from 2019 till date. Based on diagnostic modalities, the prevalence of MAFLD for ultrasound and index-based tests was 47.0% (95% CI: 31.8–62.8) and 51.2% (95% CI: 38.4–63.8) respectively. The test of subgroup difference (*X*^2^) in MAFLD prevalence did not show a significant variation. The result of the meta-regression revealed that the region, year of study publication, and diagnostic modalities did not significantly influence the overall prevalence of MAFLD among persons with T2D in Africa (**Table 2**).

**Table 2.** Subgroup and meta-regression.

| Subgroup | Studies | Prevalence<br>(95% CI) | $I^2$ (%) | P<br>value | $X^2$<br>value | P<br>value | Coefficient | P<br>value |
| --- | --- | --- | --- | --- | --- | --- | --- | --- |
| **Africa region** |  |  |  |  | 0.63 | 0.43 | -0.4211 | 0.47 |
| Sub-Saharan | 7 | 44.7 (28.7–62.0) | 96 | < 0.01 |  |  |  |  |
| North | 3 | 55.3 (36.2–73.0) | 96 | < 0.01 |  |  |  |  |
| **Publication**<br>**year** |  |  |  |  | 0.95 | 0.33 | -0.5794 | 0.26 |
| < 2019 | 5 | 40.5 (18.6–67.0) | 97 | < 0.01 |  |  |  |  |
| ≥ 2019 | 5 | 55.1 (43.6–66.1) | 93 | < 0.01 |  |  |  |  |
| **Diagnostic**<br>**modality** |  |  |  | < 0.01 | 0.16 | 0.69 | -0.1850 | 0.79 |
| Ultrasound | 8 | 47.0 (31.8–62.8) | 96 | < 0.01 |  |  |  |  |
| Index-based | 2 | 51.2 (38.4–63.8) | 81 | 0.02 |  |  |  |  |

### Publication bias

The funnel plot for studies that assessed the prevalence of MAFLD among persons with T2D showed no asymmetry, indicating that no publication bias was present (**Figure 3**). This was statistically confirmed with Egger’s test (p = 0.4754).

**Figure 3.**
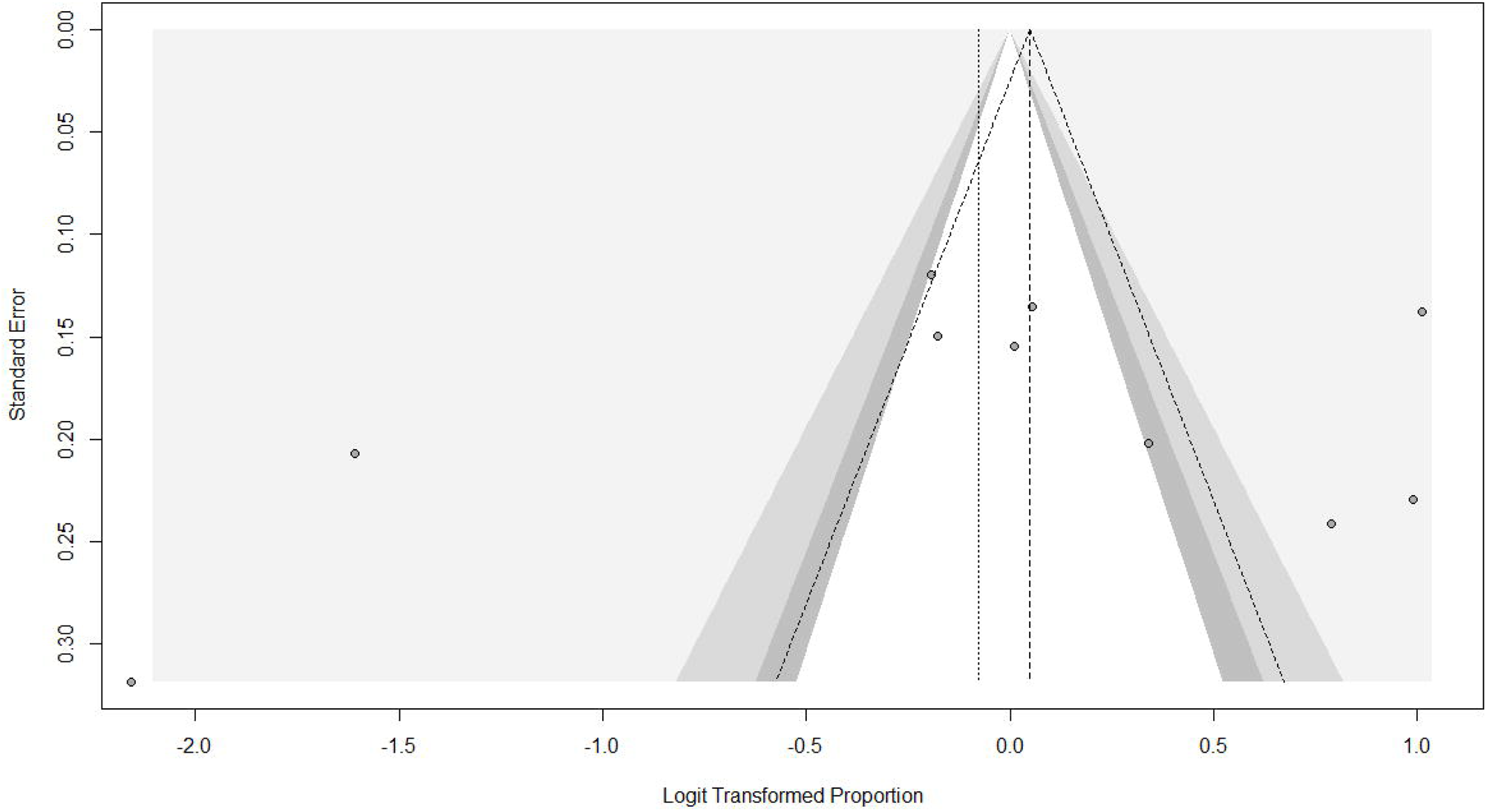
Funnel plot for MAFLD in persons with T2D.

### Association between MAFLD and glycemic control

Two articles assessed the association between MAFLD and HbA1c, with both identifying a significant positive correlation between these variables [26, 28]. The correlation coefficient recorded 0.27 in Afolabi et al.’s study [26] and 0.35 in Abebe et al.’s study [28]. As shown in **figure 4**, the result of the meta-analysis indicates a weak positive correlation between MAFLD and HbA1c (r = 0.33, 95% CI: 0.18 – 0.47).

**Figure 4.**
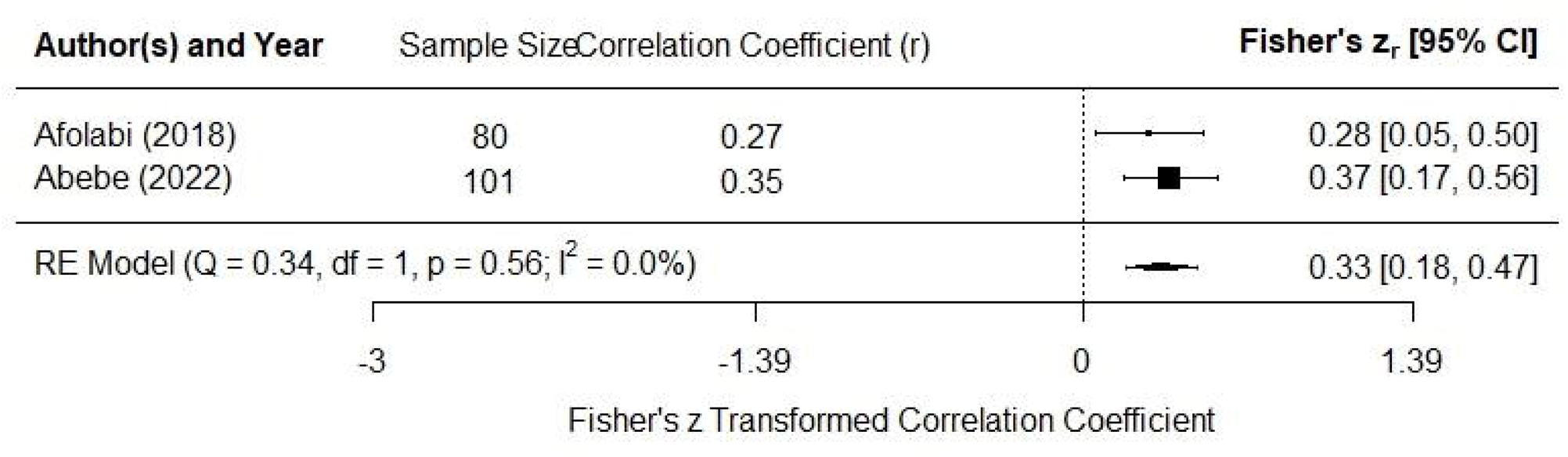
Forest plot for the association between MAFLD and glycemic control.

## Discussion

Previous global meta-analysis has identified the prevalence of MAFLD among persons with T2D in Africa to be 30.4% [7]. However, this evidence is limited in scope as the prevalence estimate was obtained through a sub-group analysis incorporating only four studies. Given the small number of studies, it is likely that the prevalence estimate may not adequately represent the diverse populations and healthcare contexts across Africa. This present systematic review and meta-analysis is the first of its kind to assess the prevalence of MAFLD among persons with T2D, with a focus on the African continent. Additionally, it is novel in exploring the association between MAFLD and glycemic control among persons with T2D in Africa.

Our meta-analysis, encompassing ten studies, revealed a substantial 48.1% prevalence of MAFLD among individuals with T2D in Africa. This prevalence was notably high in both Northern and sub-Saharan Africa, indicating that MAFLD poses a significant burden that transcends geographical barriers within the region. Interestingly, our findings surpass the prevalence of MAFLD in the general population in the same region, which was determined to be 28.2% [34]. This high prevalence is concerning and supports the American Diabetes Association’s recommendations for screening, early detection and treatment of MAFLD in persons with T2D [13].

Consistent with the study by Younossi et al., our findings suggest that the prevalence of MAFLD among individuals with T2D in Africa is comparatively lower than global and regional estimates, including Asia, Europe, and Latin America [7]. However, with the increasing adoption of Westernized lifestyle (a factor known to perpetuate MAFLD and T2D) in Africa [35], it is likely that future studies will observe an increasing trend in the prevalence rate. Notably, our subgroup analysis revealed a noticeable increase in the prevalence rate of MAFLD among persons with T2D in Africa, recording 55.1% in studies published after 2019. This indicates a changing landscape and underscores the need for continuous monitoring and intervention strategies to address the rising burden of MAFLD among individuals with T2D in the African continent.

HbA1c stands out as a strong predictor of complications associated with diabetes, as well as diabetes-related deaths. Persons with T2D and exhibiting elevated HbA1c levels face an increased risk of developing both macrovascular and microvascular complications [36, 37]. Consequently, treatment objectives for T2D frequently emphasize the importance of maintaining optimal HbA1c levels [38]. A previous study has demonstrated that improvement in glycemic control in persons with T2D was associated with improvement in MAFLD [39]. Thus, our finding that MAFLD is positively associated with HbA1c underscores the interconnected nature of these conditions and highlights the need for targeted interventions.

Although the underlying mechanism of the association between MAFLD and glycemic control is not fully understood, studies have delineated plausible pathways by which improvements in glycemic control might improve the course of MAFLD. In individuals with diabetes and MAFLD, de novo lipogenesis (DNL) significantly contributes to hepatic triglyceride accumulation. Improved glycemic control can restrict the availability of glucose as a substrate for hepatic DNL [39]. Additionally, glucose plays a role in activating stellate cells, which is pivotal in the fibrotic response of MAFLD [40]. Therefore, optimizing glycemic control has the potential to modulate the fibrotic response to lipid and inflammatory factors, key drivers of advanced MAFLD.

### Strength and limitation

The major strength of our study was the inclusion of relatively higher number of studies, providing a more comprehensive assessment of the prevalence of MAFLD among individuals with T2D in Africa. However, the findings may not be fully generalizable to all populations in Africa as the included studies were from only six countries and excluded non-English articles. Moreover, our result on the association between MAFLD and glycemic control was gleaned from only two studies. Future research endeavors should aim for more extensive geographic representation, encompassing diverse populations within the continent. Furthermore, the meta-analysis on the MAFLD prevalence exhibited a high level of heterogeneity which remained unexplained even after examining some of the potential moderators. It is worth noting that the studies incorporated in this review excluded participants with secondary liver etiologies (such excessive alcohol consumption) which does not capture the entire spectrum of MAFLD.

## Conclusions

This review identified a high prevalence of MAFLD among persons with T2D which is likely to transcend across the geographical areas of Africa. MAFLD was positively associated with glycemic control. Our findings highlight the importance of adopting a comprehensive approach to diabetes care in Africa, one that includes regular screening and intervention for MAFLD.

## Funding

No funding was received to undertake this project.

## Conflict of interest

The authors have declared that no competing interests exist.

## Data Availability Statement

All data for this review can be accessed in this manuscript and its supporting information files.

## Supporting information

**S1 Checklist. PRISMA 2009 checklist**

**S1 Text. Search strategy**

**S1 Table. Quality assessment of included studies**

## Notes

### Competing Interest Statement

The authors have declared no competing interest.

### Funding Statement

The authors received no financial support for this work

