## Supporting information for "Prevalence of metabolic dysfunction-associated fatty liver disease and its association with glycemic control in persons with type 2 diabetes in Africa: a systematic review and meta-analysis"

Quality assessment of included studies using (JBI Checklist for Prevalence Studies)

| First author (year) | Was the sample frame appropriate to address the target population? | Were study participants sampled in an appropriate way? | Was the sample size adequate? | Were the study subjects and the setting described in detail? | Was the data analysis conducted with sufficient coverage of the identified sample? | Were valid methods used for the identification of the condition? | Was the condition measured in a standard, reliable way for all participants? | Was there appropriate statistical analysis? | Was the response rate adequate, and if not, was the low response rate managed appropriately? | Score |
| --- | --- | --- | --- | --- | --- | --- | --- | --- | --- | --- |
| Zawdie (2018) | Yes | Yes | Yes | Yes | Yes | Yes | Yes | Yes | Unclear | 8 |
| Olusanya (2016) | Yes | No | Yes | Yes | Yes | Yes | Yes | Yes | Yes | 8 |
| Afolabi (2018) | Yes | No | Yes | Yes | Yes | Yes | Yes | Yes | Unclear | 7 |
| Abebe (2022) | Yes | No | Yes | Yes | Yes | Yes | Yes | Yes | Yes | 8 |
| Wiafe (2023) | Yes | Yes | Yes | Yes | Yes | Yes | Yes | Yes | Unclear | 8 |
| Assarra (2022) | Yes | Unclear | Yes | Yes | Yes | Yes | Yes | Unclear | Yes | 7 |
| El-Ashmawy (2019) | No | Unclear | Unclear | No | Yes | Yes | Yes | Yes | Unclear | 4 |
| Almobarak (2015) | Unclear | Unclear | Unclear | Yes | Yes | Yes | Yes | Unclear | Unclear | 4 |
| Fennoun (2020) | Unclear | Unclear | Unclear | Yes | Yes | Yes | Unclear | Unclear | Unclear | 3 |
| Onyekwere (2011) | Unclear | No | Unclear | Yes | Yes | Yes | Yes | Unclear | Unclear | 4 |
