## Supporting information for "Prevalence of metabolic dysfunction-associated fatty liver disease and its association with glycemic control in persons with type 2 diabetes in Africa: a systematic review and meta-analysis"

Search strategy used for the various databases

**PubMed**

| Concept | Seach string | Records |
| --- | --- | --- |
| #1 Non-alcoholic fatty liver | "Non-alcoholic Fatty Liver Disease"[Mesh] OR "Fatty Liver"[Mesh] OR "metabolic dysfunction-associated fatty liver" OR "non-alcoholic fatty liver disease" OR “nonalcoholic fatty liver disease" OR “non-alcoholic fatty liver” OR “nonalcoholic fatty liver” OR “fatty liver” OR "non‐alcoholic steatohepatitis" OR "nonalcoholic steatohepatitis" OR MAFLD OR NAFLD OR NASH | 82115 |
| #2 Type 2 diabetes | "Diabetes Mellitus"[Mesh] OR "Diabetes Mellitus, Type 2"[Mesh] OR "type 2 diabetes" OR "non-insulin-dependent diabetes mellitus" OR NIDD OR T2D OR T2DM | 568821 |
| #3 Africa | "Africa"[Mesh] OR Africa OR Algeria OR Angola OR Benin OR Botswana OR "Burkina Faso" OR Burundi OR "Cabo Verde" OR Cameroon OR "Central African Republic" OR Chad OR Comoros OR Congo OR "Democratic Republic of Congo" OR "Cote d'Ivoire" OR Djibouti OR Egypt OR "Equatorial Guinea" OR Eritrea OR Eswatini OR Ethiopia OR Gabon OR Gambia OR Ghana OR Guinea OR Guinea-Bissau OR Kenya OR Lesotho OR Liberia OR Libya OR Madagascar OR Malawi OR Mali OR Mauritania OR Mauritius OR Morocco OR Mozambique OR Namibia OR Niger OR Nigeria OR Rwanda OR "Sao Tome and Principe" OR Senegal OR Seychelles OR "Sierra Leone" OR Somalia OR "South Africa" OR "South Sudan" OR Sudan OR Tanzania OR Togo OR Tunisia OR Uganda OR Zambia OR Zimbabwe | 945723 |
| **#4** | #1 AND #2 AND #3 | 151 |

**Medline**

| Concept | Seach string | Records |
| --- | --- | --- |
| **#1** Non-alcoholic fatty liver | exp Non-alcoholic Fatty Liver Disease/ or exp Fatty Liver/ or ("metabolic dysfunction-associated fatty liver" or "non-alcoholic fatty liver disease" or "nonalcoholic fatty liver disease" or "non-alcoholic fatty liver" or "nonalcoholic fatty liver" or "fatty liver" or "non‐alcoholic steatohepatitis" or "nonalcoholic steatohepatitis" or MAFLD or NAFLD or NASH).mp. | 69891 |
| **#2** Type 2 diabetes | exp Diabetes Mellitus/ or exp Diabetes Mellitus, Type 2/ or ("type 2 diabetes" or "non-insulin-dependent diabetes mellitus" or NIDD or T2D or T2DM).mp. | 569146 |
| **#3** Africa | exp Africa/ or (Africa or Algeria or Angola or Benin or Botswana or "Burkina Faso" or Burundi or "Cabo Verde" or Cameroon or "Central African Republic" or Chad or Comoros or "Democratic Republic of the Congo" or "Republic of the Congo" or "Cote d'Ivoire" or Djibouti or Egypt or "Equatorial Guinea" or Eritrea or Eswatini or Ethiopia or Gabon or Gambia or Ghana or Guinea or Guinea-Bissau or Kenya or Lesotho or Liberia or Libya or Madagascar or Malawi or Mali or Mauritania or Mauritius or Morocco or Mozambique or Namibia or Niger or Nigeria or Rwanda or "Sao Tome and Principe" or Senegal or Seychelles or "Sierra Leone" or Somalia or "South Africa" or "South Sudan" or Sudan or Tanzania or Togo or Tunisia or Uganda or Zambia or Zimbabwe).mp. | 635221 |
| **#4** | #1 AND #2 AND #3 | 51 |

**Embase**

| Concept | Seach string | Records |
| --- | --- | --- |
| **#1** Non-alcoholic fatty liver | exp nonalcoholic fatty liver/ or exp fatty liver/ or ("metabolic dysfunction-associated fatty liver" or "non-alcoholic fatty liver disease" or "nonalcoholic fatty liver disease" or "non-alcoholic fatty liver" or "nonalcoholic fatty liver" or "fatty liver" or "non‐alcoholic steatohepatitis" or "nonalcoholic steatohepatitis" or MAFLD or NAFLD or NASH).mp. | 132012 |
| **#2** Type 2 diabetes | exp diabetes mellitus/ or exp Diabetes Mellitus, Type 2/ or ("type 2 diabetes" or "non-insulin-dependent diabetes mellitus" or NIDD or T2D or T2DM).mp. | 1276641 |
| **#3** Africa | exp Africa/ or (Africa or Algeria or Angola or Benin or Botswana or "Burkina Faso" or Burundi or "Cabo Verde" or Cameroon or "Central African Republic" or Chad or Comoros or "Democratic Republic of the Congo" or "Republic of the Congo" or "Cote d'Ivoire" or Djibouti or Egypt or "Equatorial Guinea" or Eritrea or Eswatini or Ethiopia or Gabon or Gambia or Ghana or Guinea or Guinea-Bissau or Kenya or Lesotho or Liberia or Libya or Madagascar or Malawi or Mali or Mauritania or Mauritius or Morocco or Mozambique or Namibia or Niger or Nigeria or Rwanda or "Sao Tome and Principe" or Senegal or Seychelles or "Sierra Leone" or Somalia or "South Africa" or "South Sudan" or Sudan or Tanzania or Togo or Tunisia or Uganda or Zambia or Zimbabwe).mp. | 674259 |
| **#4** | #1 AND #2 AND #3 | 182 |

**Global Health**

| Concept | Seach string | Records |
| --- | --- | --- |
| **#1** Non-alcoholic fatty liver | exp non-alcoholic fatty liver/ or exp fatty liver/ or ("metabolic dysfunction-associated fatty liver" or "non-alcoholic fatty liver disease" or "nonalcoholic fatty liver disease" or "non-alcoholic fatty liver" or "nonalcoholic fatty liver" or "fatty liver" or "non‐alcoholic steatohepatitis" or "nonalcoholic steatohepatitis" or MAFLD or NAFLD or NASH).mp. | 20258 |
| **#2** Type 2 diabetes | exp diabetes/ or exp type 2 diabetes/ or ("type 2 diabetes" or "non-insulin-dependent diabetes mellitus" or NIDD or T2D or T2DM).mp. | 141796 |
| **#3** Africa | exp Africa/ or (Africa or Algeria or Angola or Benin or Botswana or "Burkina Faso" or Burundi or "Cabo Verde" or Cameroon or "Central African Republic" or Chad or Comoros or "Democratic Republic of the Congo" or "Republic of the Congo" or "Cote d'Ivoire" or Djibouti or Egypt or "Equatorial Guinea" or Eritrea or Eswatini or Ethiopia or Gabon or Gambia or Ghana or Guinea or Guinea-Bissau or Kenya or Lesotho or Liberia or Libya or Madagascar or Malawi or Mali or Mauritania or Mauritius or Morocco or Mozambique or Namibia or Niger or Nigeria or Rwanda or "Sao Tome and Principe" or Senegal or Seychelles or "Sierra Leone" or Somalia or "South Africa" or "South Sudan" or Sudan or Tanzania or Togo or Tunisia or Uganda or Zambia or Zimbabwe).mp. | 398834 |
| **#4** | #1 AND #2 AND #3 | 39 |

**Scopus**

| Concept | Seach string | Records |
| --- | --- | --- |
| **#1** Non-alcoholic fatty liver | TITLE-ABS-KEY ("metabolic dysfunction-associated fatty liver" OR "non-alcoholic fatty liver disease" OR "nonalcoholic fatty liver disease" OR "non-alcoholic fatty liver" OR "nonalcoholic fatty liver" OR "fatty liver" OR "non‐alcoholic steatohepatitis" OR "nonalcoholic steatohepatitis" OR MAFLD OR NAFLD OR NASH) | 140985 |
| **#2** Type 2 diabetes | TITLE-ABS-KEY (Diabetes OR "type 2 diabetes" OR "non-insulin-dependent diabetes mellitus" OR NIDD OR T2D OR T2DM) | 1155227 |
| **#3** Africa | TITLE-ABS-KEY (Africa OR Algeria OR Angola OR Benin OR Botswana OR "Burkina Faso" OR Burundi OR "Cabo Verde" OR Cameroon OR "Central African Republic" OR Chad OR Comoros OR "Democratic Republic of the Congo" OR "Republic of the Congo" OR "Cote d'Ivoire" OR Djibouti OR Egypt OR "Equatorial Guinea" OR Eritrea OR Eswatini OR Ethiopia OR Gabon OR Gambia OR Ghana OR Guinea OR "Guinea-Bissau" OR Kenya OR Lesotho OR Liberia OR Libya OR Madagascar OR Malawi OR Mali OR Mauritania OR Mauritius OR Morocco OR Mozambique OR Namibia OR Niger OR Nigeria OR Rwanda OR "Sao Tome and Principe" OR Senegal OR Seychelles OR "Sierra Leone" OR Somalia OR "South Africa" OR "South Sudan" OR Sudan OR Tanzania OR Togo OR Tunisia OR Uganda OR Zambia OR Zimbabwe) | 1425983 |
| **#4** | #1 AND #2 AND #3 | 166 |

**Web of Science**

| Concept | Seach string | Records |
| --- | --- | --- |
| **#1** Non-alcoholic fatty liver | Topic ("metabolic dysfunction-associated fatty liver" OR "non-alcoholic fatty liver disease" OR "nonalcoholic fatty liver disease" OR "non-alcoholic fatty liver" OR "nonalcoholic fatty liver" OR "fatty liver" OR "non‐alcoholic steatohepatitis" OR "nonalcoholic steatohepatitis" OR MAFLD OR NAFLD OR NASH) | 113242 |
| **#2** Type 2 diabetes | Topic (Diabetes OR "type 2 diabetes" OR "non-insulin-dependent diabetes mellitus" OR NIDD OR T2D OR T2DM) | 765299 |
| **#3** Africa | Topic (Africa OR Algeria OR Angola OR Benin OR Botswana OR "Burkina Faso" OR Burundi OR "Cabo Verde" OR Cameroon OR "Central African Republic" OR Chad OR Comoros OR "Democratic Republic of the Congo" OR "Republic of the Congo" OR "Cote d'Ivoire" OR Djibouti OR Egypt OR "Equatorial Guinea" OR Eritrea OR Eswatini OR Ethiopia OR Gabon OR Gambia OR Ghana OR Guinea OR "Guinea-Bissau" OR Kenya OR Lesotho OR Liberia OR Libya OR Madagascar OR Malawi OR Mali OR Mauritania OR Mauritius OR Morocco OR Mozambique OR Namibia OR Niger OR Nigeria OR Rwanda OR "Sao Tome and Principe" OR Senegal OR Seychelles OR "Sierra Leone" OR Somalia OR "South Africa" OR "South Sudan" OR Sudan OR Tanzania OR Togo OR Tunisia OR Uganda OR Zambia OR Zimbabwe) | 1057470 |
| **#4** | #1 AND #2 AND #3 | 63 |
